# The impact of the COVID-19 lockdown on maternal mental health and coping in the UK: Data from the COVID-19 New Mum Study

**DOI:** 10.1101/2020.08.04.20168039

**Authors:** Sarah Dib, Emeline Rougeaux, Adriana Vázquez, Jonathan Wells, Mary Fewtrell

## Abstract

**Background:** Depression and anxiety affect up to 20% of new and expectant mothers during the perinatal period; this rate may have increased due to COVID-19 and lockdown measures. This analysis aimed to assess how mothers are feeling and coping during lockdown, and to identify the potential pathways that can assist them.

**Methods:** 1329 women living in the UK aged ≥18 years with an infant ≤12 months of age completed an anonymous online survey. Descriptive analysis of maternal mental health, coping, support received, activities undertaken and consequences of lockdown was conducted. Linear regression was used to predict maternal mental health and coping, using activities, support, and consequences of the lockdown as predictors, while adjusting for age, gestational age, ethnicity, income, marital status and number of children.

**Results:** More than half of the participants reported feeling down (56%), lonely (59%), irritable (62%) and worried (71%), to some or high extent since lockdown began. Despite this, 70% felt able to cope with the situation. Support with her own health (95% CI .004,235), contacting infant support groups (95% CI -.003, .252), and higher infant gestational age (95% CI .000, .063) predicted better mental health. Travelling for work (95% CI -.680, - .121), lockdown having a major impact on the ability to afford food (95% CI -1.202, -.177), and having an income lower than 30k (95% CI -.475, -.042) predicted poorer mental health. Support with her own health and more equal division of household chores were associated with better coping.

**Conclusion:** During lockdown, a large proportion of new mothers experienced symptoms of poor mental health; mothers of infants with lower gestational age, with low income, and who are travelling to work were particularly at risk. However, greater support for maternal health and with household chores showed positive associations with maternal mental health and coping. These findings highlight the urgent need to assess maternal mental health, and to identify prevention strategies for mothers during different stages of lockdown.

## Background

The coronavirus disease 2019 (COVID-19) pandemic resulted in a rapid change of circumstances in the UK population, and many individuals may have experienced loss of livelihood, increased financial burden, reduced personal support systems and professional services, physical isolation, and illness. Therefore, mental health is likely to be affected, as is already evident. A recent Lancet paper has highlighted the urgent need for research that tackles how the effects of COVID-19 on mental health, especially in vulnerable populations, could be eased [1].

These effects may be of particular importance for mothers during pregnancy and in the first year after giving birth. According to data from before the pandemic, perinatal mental illness affects up to 20% of new and expectant mothers, and is associated with increased risk of preterm delivery, reduced mother-infant bonding and decreased odds of breastfeeding [2, 3]. It is expected that this rate may have increased [4] as a result of physical and social isolation, changes in perinatal services, and the economic burden of the disease that is disproportionately affecting women [5, 6]. Several studies have highlighted the increase in distress and psychological problems experienced by pregnant women during the pandemic [7-9], but limited evidence has been published regarding postnatal mental health. A recent study from Canada has shown that there is a substantial increase in self-reported depression and anxiety in pregnant and postpartum women [10]. In the UK, we recently reported that new mothers with babies under 1 year of age expressed feelings of being robbed of the joys of motherhood [11], which was also noted in a recent qualitative study [12].

The pandemic has further highlighted health and social inequalities, where Black, Asian and minority ethnic (BAME) groups, people with disabilities or pre-existing health conditions, and socioeconomically disadvantaged groups have been disproportionately impacted [13, 14]. Recent data has shown the higher risk of death and serious illness due to COVID-19 in these groups. Therefore, extra attention should be given to mothers who belong to these groups to prevent or manage perinatal distress and mitigate inequalities.

The COVID-19 New Mum Study aims to capture information on maternal experiences, mood and infant feeding practices during the different phases of lockdown. This will help in understanding the experiences and needs of new mothers, and ultimately in improving support and advice given. We have previously highlighted reductions in support for infant feeding, childcare support, and own health, in addition to changes in birth and infant feeding plans [11]. In this report, we present descriptive data on maternal mental health, coping, support and activities during week 1 of the study (May 27th-June 3rd, 2020). We also investigate possible pathways for the effects of COVID-19 lockdown on maternal mental health and coping.

## Methods

### Study Design

Women living in the UK aged ≥18 years who have an infant currently under 12 months of age are being invited to complete a one-time, anonymous online survey (The Covid-19 New Mum Survey – https://is.gd/covid19newmumstudy). The survey was launched on May 27^th^ and will remain open until December 31^st^ 2020 to capture data relating to the different levels of lockdown restrictions.

The survey is being advertised mainly on social media groups and pages used by mothers such as Facebook infant feeding and mother and baby support groups, Twitter, and Instagram. Information about the study is also being disseminated via contacts with relevant professional and support organisations and groups and via word of mouth.

### Survey

The full content of the survey that includes details about the background factors, infant feeding practices, impacts of COVID-19 has been described elsewhere [11]. For this analysis, we collected data regarding:

#### 1. Background characteristics

Social and demographic factors such as maternal age and ethnicity, maternal education and household income, infant‘s age and gestational age, and living conditions (access to green space, type of accommodation).

#### 2. Consequences of lockdown

whether they have been advised to shield/isolate, and whether they gave birth before or during lockdown. We also asked about the impact of lockdown on the family‘s ability to afford rent, food, and other essentials.

#### 3. Mother‘s activities, access to support and perceptions

Questions ask about how often (0 times, 1 to 3 times, 4 or 5 times, or daily) the participants engaged in activities in the previous week, such as walking, exercise, relaxation techniques and grocery shopping. Frequency of accessing infant support groups and contact with health care and mental health professionals was also collected to assess the level of support received. Maternal perceptions of lockdown were assessed retrospectively where participants were asked to state how much the following statements applied to them since lockdown began (not at all, very little, to some extent, to a high extent): I‘ve been feeling down, I‘ve been feeling lonely, I‘ve had trouble relaxing, I‘ve become easily annoyed or irritable and I‘ve been feeling worried. Appetite and sleep disruption was also similarly assessed. We also included ‘positive’ statements, these include: I feel able to cope with the situation, I‘ve enjoyed the spring weather, I‘ve had the opportunity to chat with my family and friends, I‘ve had time to enjoy personal interests or hobbies, I‘ve had time to focus on my health and I‘ve had time to exercise.

Care was taken to minimise or avoid questions that might provoke or worsen psychological distress. Additionally, since this is an anonymous survey and the participants cannot be identified for follow up, no formal assessment of depression or anxiety (using validated tools) was undertaken. Participants were given the option to omit any sensitive questions they did not wish to answer. A list of resources for infant feeding and maternal mental health support, including resources from organisations providing services specifically to people belonging to BAME, LGBTQ+ and disabled groups, is provided at the end of the survey.

### Ethics and Consent

Ethical approval was obtained from the UCL Research Ethics committee (0326/017). The first page of the survey provides information about the study and, having read this, participants were asked to provide consent to participate before proceeding.

### Statistical Analysis

Data from the survey was exported from RedCap, the software collecting survey responses, and analysed in SPSS (IBM SPSS statistics v. 26). Descriptive data are shown as mean (SD) or n (%). Descriptive characteristics of the population, and description of maternal mental health, support received, activities undertaken, and consequences of lockdown were presented for the whole sample. Principal Component Analysis (PCA) was conducted for the maternal perceptions; Kaiser-Meyer-Olkin Measure of Sampling Adequacy and Bartlett‘s test of sphericity were used to assess whether PCA is appropriate. Eigenvalues (of 1), Scree plots and parallel analysis were used to identify the components of perceptions. Regression variables were saved and then linear regression was conducted to assess the relationship between mental health and coping (outcomes) and support, activities, and consequences of lockdown (as predictors). Directed acyclic graphs (DAG), as shown in Figure 1, were sketched to identify the minimum adjustment set of confounders, and to identify ancestors of the outcome that were dropped from the regression model. On this basis, the linear regression models were controlled for maternal age, baby‘s age, ethnicity, gestational age, income, marital status, and number of children in the household. 95% Confidence Intervals (CI) are presented for all regression coefficients, and p-values under 0.05 are considered statistically significant.

**Figure 1.**
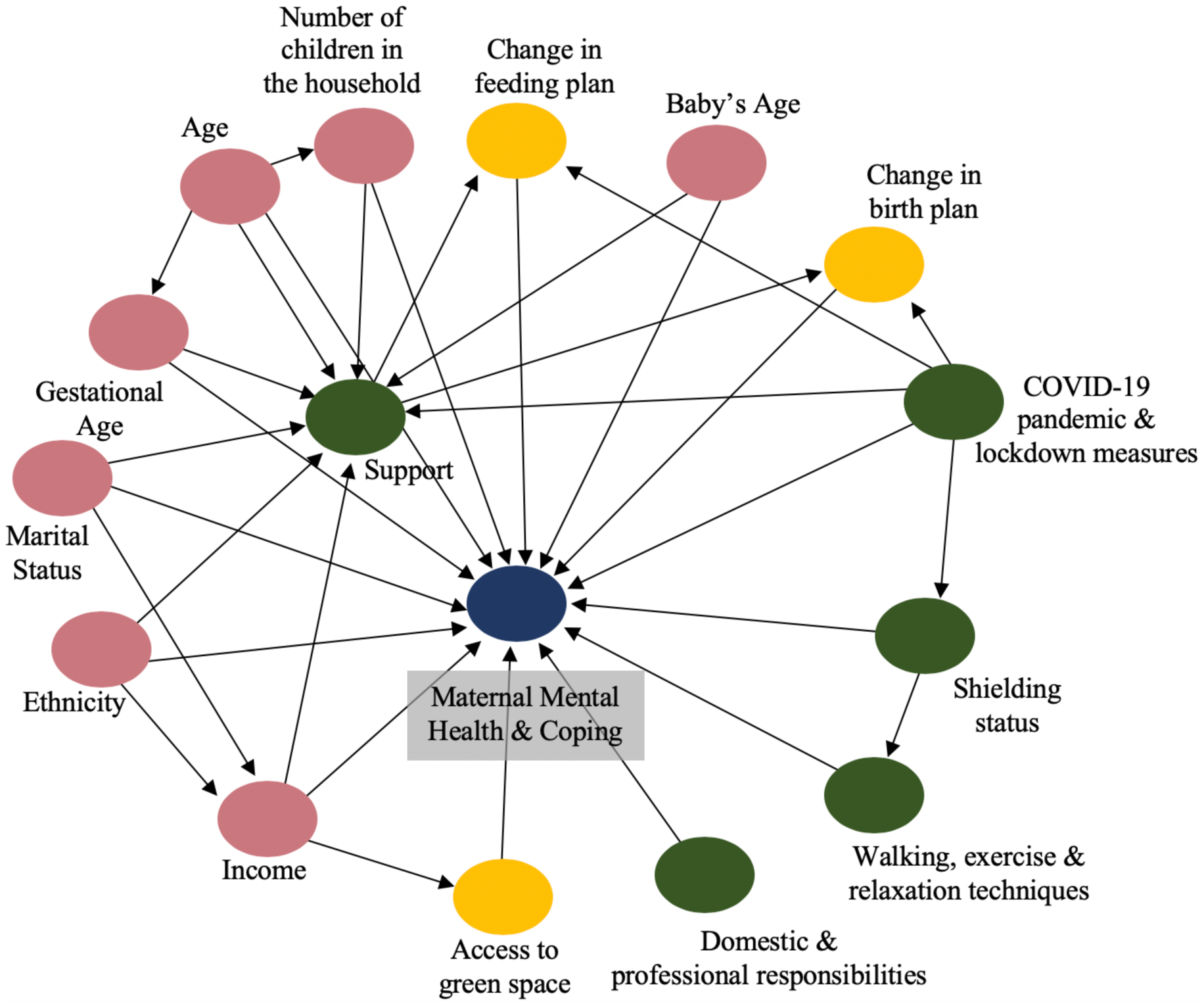
Directed acyclic graph of the relationship between maternal mental health or coping and activities, support and consequences of lockdown. It also exhibits the confounders of the relationship (such as age, marital status, income, ethnicity). Ancestors of the outcome are then dropped from the regression model. The blue circle represents the outcome, the green represents the predictors, the pink represent the covariates and the yellow represents the ancestor of the outcome.

## Results

### 1. Descriptive Data

#### Background Characteristics

During the first week of the study, 1329 participants fully completed the survey providing data on background characteristics, mental health, support, consequences of lockdown, and coping mechanisms. Background characteristics are presented in Table 1. The majority of participants self-identified as white, were married or living with a partner, lived in a house, and had access to green space within walking distance.

**Table 1.**
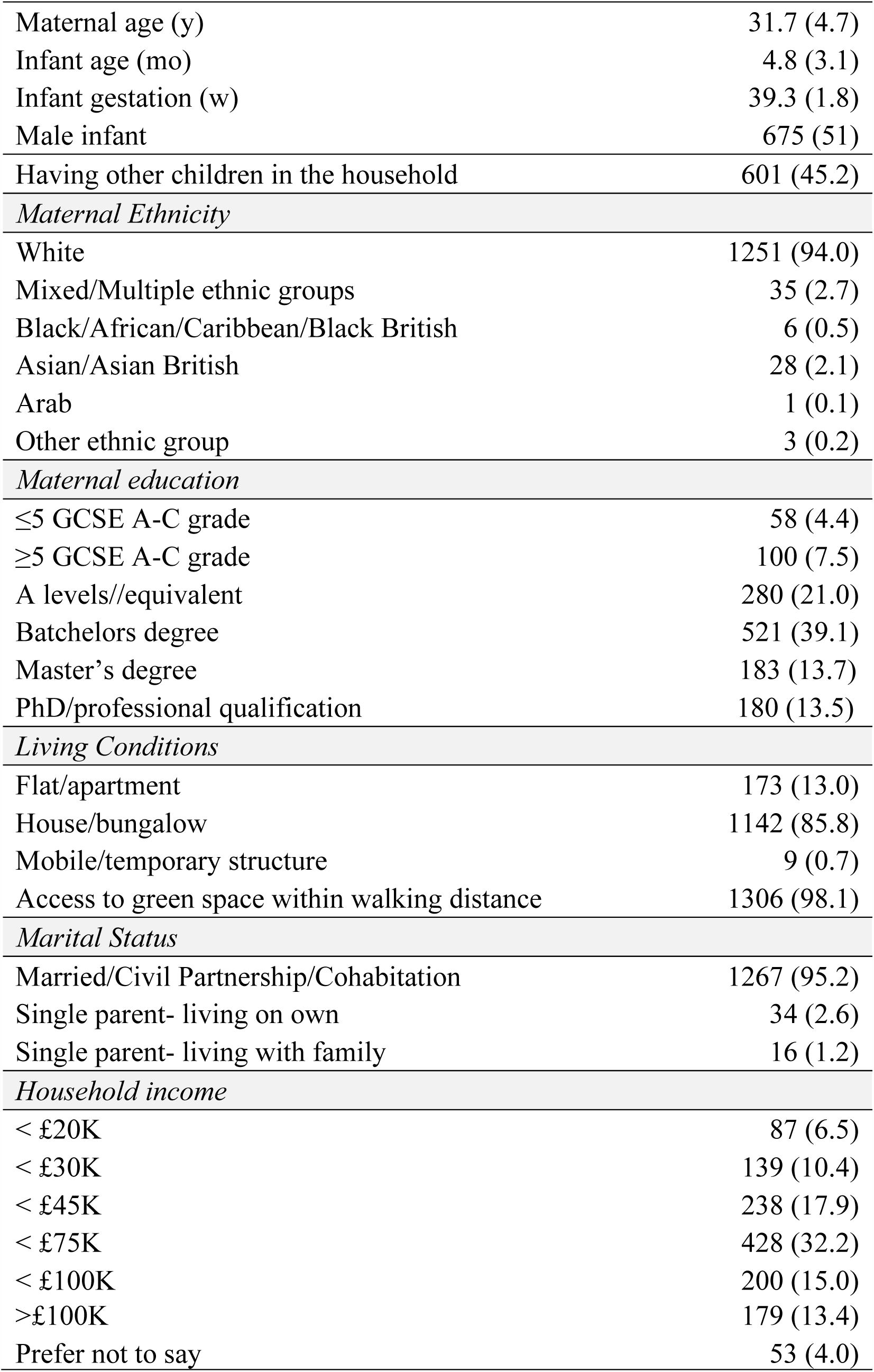
Background characteristics of women who completed the survey in week 1 (mean (SD) or n(%))

#### Perceptions of lockdown: mental health, time availability, coping, appetite and sleep

PCA was conducted for the questions pertaining to perceptions during lockdown. Four components were identified, which were labelled as follows: 1) ‘maternal mental health’ (an indicator of low mood, anxiety and loneliness, although not formally assessed), 2) ‘time availability’ to exercise, enjoy personal interests and focus on health, 3) ‘coping’ 4) appetite and sleep changes (Supplementary Table 1). The four components and corresponding perceptions are presented in Table 2. In this paper, we focus on the maternal mental health and coping components as outcomes.

**Table 2.** The extent to which the participants agreed with the following perceptions, since the lockdown began n (%).

|  |  | Not at all | Very little | To some extent | To a high extent |
| --- | --- | --- | --- | --- | --- |
| *Maternal mental health* | I've been feeling down | 161 (12.1) | 414 (31.1) | 529 (39.7) | 222 (16.7) |
|  | I've been feeling lonely | 232 (17.4) | 310 (23.3) | 472 (35.5) | 316 (23.7) |
|  | I've had trouble relaxing | 220 (16.5) | 336 (25.2) | 465 (34.9) | 301 (22.6) |
|  | I've become easily annoyed or irritable | 143 (10.7) | 354 (26.6) | 502 (37.7) | 328 (24.6) |
|  | I've been feeling worried | 98 (7.4) | 291 (21.9) | 532 (40.0) | 407 (30.6) |
| *Coping* | I've had the opportunity to chat with my family and friends | 4 (0.3) | 100 (7.5) | 602 (45.2) | 619 (46.5) |
|  | I've enjoyed the spring weather | 46 (3.5) | 150 (11.3) | 485 (36.4) | 645 (48.5) |
|  | I feel able to cope with the situation | 85 (6.4) | 320 (24.0) | 748 (56.2) | 178 (13.4) |
| *Appetite and sleep changes* | I've had trouble falling or staying asleep | 460 (34.6) | 339 (25.5) | 326 (24.5) | 205 (15.4) |
|  | I've been having poor appetite | 848 (63.7) | 230 (17.3) | 196 (14.7) | 49 (3.7) |
| *Time availability to focus on health and interests* | I've had time to enjoy personal interests | 635 (47.7) | 476 (35.8) | 169 (12.7) | 48 (3.6) |
|  | I've had time to focus on my health | 374 (28.1) | 549 (41.2) | 313 (23.5) | 93 (7.0) |
|  | I've had time to exercise | 284 (21.3) | 481 (36.1) | 391 (29.4) | 173 (13.0) |

As shown in Table 2, more than half of the participants reported feeling down, lonely and irritable, and had trouble relaxing, to some or to a high extent. Seventy-one percent of mothers expressed feeling worried to some or to a high extent since lockdown began.

Forty percent and 18% of participants reported having trouble falling asleep and poor appetite, respectively, to some or to a high extent.

Additionally, 83% and 60% of mothers stated having no or very little time to enjoy personal interests or to exercise, respectively. Seventy percent said they had no or very little time to focus on their own health.

Despite these perceptions, the majority of the mothers expressed feeling able to cope with the situation (70%) and enjoy the weather (85%), and having the opportunity to connect with family and friends (92%) to some or to a high extent.

#### Activities during lockdown

Table 3 shows the activities the participants engaged in in the previous week. While 95% of participants did not travel for work in the previous week, 95% went out for a walk or exercise, 53% went shopping at the grocery or pharmacy, and 23% practiced a relaxation technique at least once a week. The most commonly cited relaxation techniques were yoga, meditation, and breathing; 37% of mothers started using these methods during lockdown whereas the rest practiced relaxation techniques regularly before the lockdown.

**Table 3.** Activities the participants engaged in in the previous week n (%)

|  | 0 times | 1 to 3 times | 4 or 5 times | Daily or more |
| --- | --- | --- | --- | --- |
| Went shopping at the grocery store or pharmacy | 625 (47.2) | 670 (50.6) | 25 (1.9) | 4 (0.3) |
| Travelled for work | 1269 (95.3) | 44 (3.3) | 4 (0.3) | 7 (0.5) |
| Went outside for a walk or for exercise | 67 (5.1) | 438 (33.2) | 296 (22.4) | 520 (39.4) |
| Practiced a relaxation technique | 1021 (76.7) | 218 (16.4) | 57 (4.3) | 32 (2.4) |

Fifty-five mothers travelled to work at least once a week. Of these, 38% (n=21) worked in the NHS or medical/healthcare sector, followed by business/administration/accounting (n=9) then education (n=7).

#### Support during lockdown

The different types of support received were assessed as shown in Table 4. Fifty-nine percent of mothers reported that they received or were receiving enough support with their own health, whereas 41% did not perceive this to be the case. Sixty-three percent did not feel, or felt only a little, that the household chores were more equally divided among household members since the lockdown began. Moreover, 27%, 41% and 7% had contact at least once in the previous week with a mother and baby or breastfeeding support group, health care professional (such as GP, health visitor, midwife) and mental health professional, respectively. Only 9 participants reported attending a face-to-face mother and baby or breastfeeding support group and 4 had an in-person session with a mental health professional, whereas the rest had contact online or by phone.

**Table 4.** Support measures for women who completed the survey in week 1 n (%)

|  |  |
| --- | --- |
| *Got or getting enough support and help with own health* |  |
| Yes | 783 (58.8) |
| No | 545 (40.9) |
| *Had contact with a mother and baby or breastfeeding support group\** |  |
| 0 times | 976 (73.3) |
| 1 to 3 times | 238 (17.9) |
| 4 or 5 times | 53 (4.0) |
| Daily or more | 58 (4.4) |
| *Had contact with a health professional (GP, health visitor, midwife)\** |  |
| 0 times | 791 (59.4) |
| 1 to 3 times | 496 (37.3) |
| 4 or 5 times | 31 (2.3) |
| Daily or more | 5 (0.4) |
| *Attended an appointment with a mental health professional\** |  |
| 0 times | 1236 (92.9) |
| 1 to 3 times | 81 (6.1) |
| 4 or 5 times | 10 (0.8) |
| Daily or more | 1 (0.1) |
| *I feel the house chores are more equally divided among household members* |  |
| Not at all | 494 (37.1) |
| Very little | 343 (25.8) |
| To some extent | 298 (22.4) |
| To a high extent | 186 (14.0) |

\*in the previous week

#### Consequences of lockdown

Seventy-three percent of mothers delivered before the lockdown started, and the rest gave birth during the lockdown period which we have previously shown had implications for birth and feeding plans. Ninety-two percent of mothers were not advised by the NHS to stay at home due to a pre-existing health condition. Thirty-seven percent expressed a minor to major impact of lockdown on the ability to pay rent or mortgage payments, 32% on ability to pay for food, and 28% on ability to pay for other essentials such as utilities and medication. The rest reported no impact.

### 2. Predictors of Maternal Mental Health and Coping

#### Predictors of maternal mental health (low mood, anxiety symptoms and loneliness)

Results of the multiple linear regression model including the predictors and confounders of maternal mental health are displayed in Table 5. Getting enough support with her own health and contact with mother and baby support groups were predictors of better mental health.

**Table 5.** Predictors of maternal mental health (anxiety, low mood and loneliness)

|  | Variable | B | 95% CI |  | p-value |
| --- | --- | --- | --- | --- | --- |
| *In person, by phone or online contact with:* | Health care professionals<sup>g</sup> | -.045 | -.167 | .077 | .470 |
|  | **Mother & baby support groups<sup>g</sup>** | **.124** | **-.003** | **.252** | **.056** |
|  | Mental health professional<sup>g</sup> | -.197 | -.418 | .025 | .082 |
|  | **Enough help with own health** | **.120** | **.004** | **.235** | **.042** |
| *Extent to which chores are more equally divided<sup>b</sup>* | Not at all | .101 | -.073 | .274 | .255 |
|  | **Very little** | **.256** | **.075** | **.438** | **.006** |
|  | To some extent | .091 | -.094 | .277 | .335 |
|  | Shopped at grocery store/pharmacy<sup>g</sup> | .063 | -.051 | .176 | .278 |
|  | Went outside for a walk or exercise<sup>g</sup> | .138 | -.109 | .385 | .274 |
|  | **Travelled for work<sup>g</sup>** | **-.401** | **-.680** | **-.121** | **.005** |
|  | Practiced a relaxation technique<sup>g</sup> | .113 | -.020 | .247 | .095 |
|  | Gave birth during lockdown | .086 | -.082 | .254 | .313 |
|  | Advised to shield due to risk | -.045 | -.318 | .228 | .745 |
|  | Minor | .075 | -.101 | .251 | .405 |
| *Impact of lockdown on the ability to afford rent<sup>c</sup>* | Moderate | .117 | -.110 | .345 | .312 |
|  | Major | .063 | -.312 | .437 | .743 |
|  | Minor | .170 | -.035 | .375 | .105 |
| *Impact of lockdown on the ability to afford food<sup>c</sup>* | Moderate | -.204 | -.518 | .109 | .202 |
|  | **Major** | **-.689** | **-1.202** | **-.177** | **.008** |
| *Impact of lockdown on the ability to afford other essentials<sup>c</sup>* | Minor | -.081 | -.313 | .151 | .494 |
|  | Moderate | .193 | -.140 | .526 | .255 |
|  | Major | .329 | -.234 | .891 | .252 |
| *Age* | Age | .004 | -.009 | .017 | .525 |
|  | Baby's age | .017 | -.007 | .041 | .154 |
|  | **Baby's gestational age** | **.031** | **.000** | **.063** | **.051** |
| *Ethnicity<sup>d</sup>* | BAME | -.166 | -.398 | .066 | .160 |
| *Marital Status<sup>e</sup>* | Single- living on own | .047 | -.311 | .405 | .798 |
|  | Single- living with family | -.078 | -.598 | .443 | .770 |
|  | **< 20-30K** | **-.258** | **-.475** | **-.042** | **.019** |
| *Income<sup>f</sup>* | < 45-75K | -.045 | -.204 | .113 | .575 |
|  | < 100K | -.067 | -.260 | .126 | .496 |
| *Number of children in the household:* | One<sup>h</sup> | -.005 | -.120 | .111 | .939 |

<sup>a</sup>PCA result of mental health. Higher scores reflect better mental health. <sup>b</sup>High extent as reference. <sup>c</sup>No impact as reference. <sup>d</sup>BAME: Black, Asian and minority ethnic group; white ethnicity as reference. <sup>e</sup>Married/civil partnership/cohabitation as reference, <sup>f</sup>Income above 100K as reference. <sup>g</sup>Participated in activities at least once in the previous week. <sup>h</sup>More than one child as reference.

Unexpectedly, mothers who reported the household chores were ‘only a little’ more equally divided had better mental health then those who reported they were more equally divided ‘to a great extent‘. Travelling for work and COVID-19 lockdown having a major impact on the ability to afford food were associated with worse mental health outcomes. A lower household income (<20-30K) also predicted poorer mental health outcomes.

#### Predictors of coping with lockdown

Results of the multiple linear regression model are presented in Table 6. Perception of getting enough support with her own health predicted better coping with the situation. Conversely, with each unit decrease in the extent to which the participants perceived the household chores have become more equally divided, coping decreases.

**Table 6.**
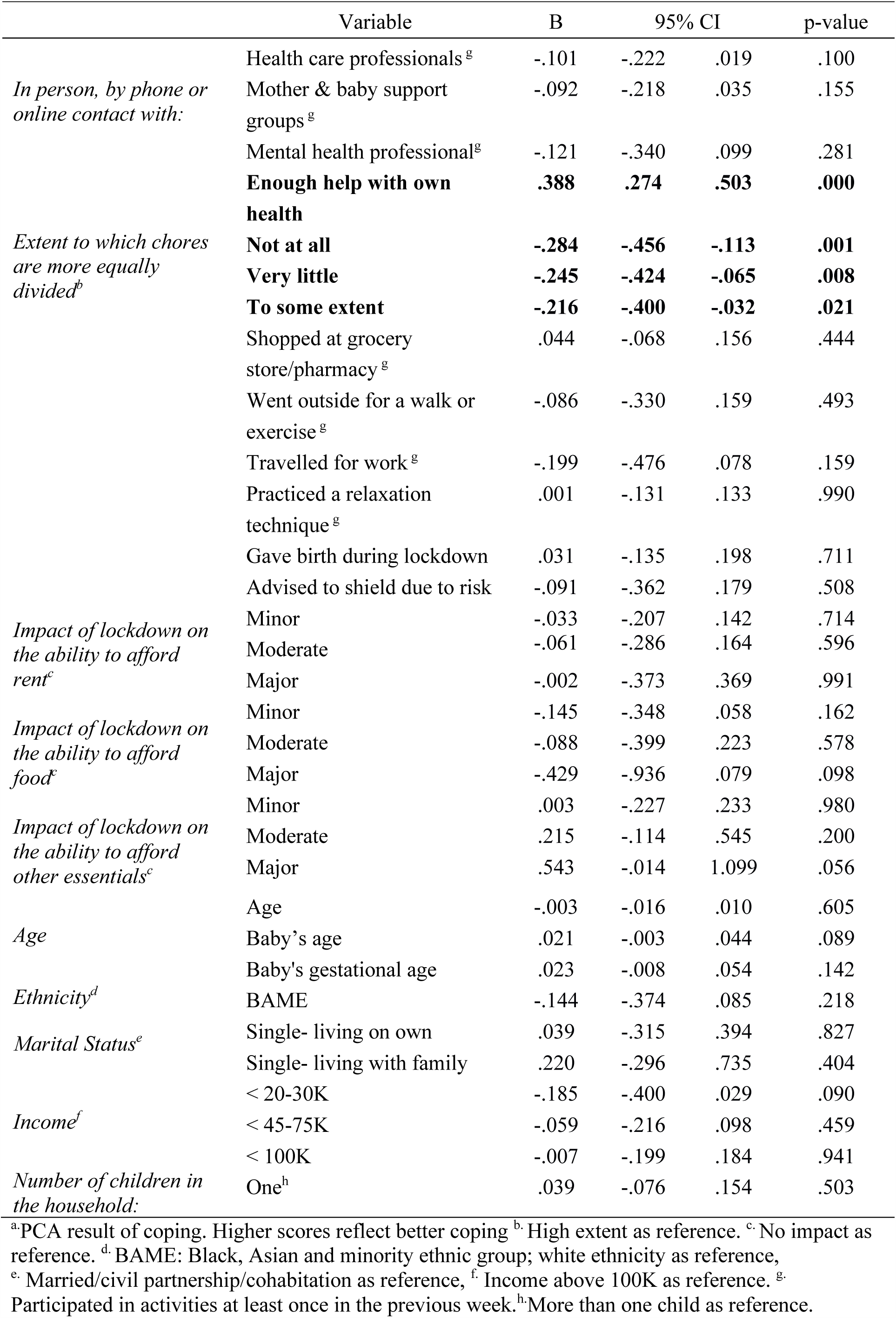
Predictors of coping

## Discussion

The findings of this survey illustrate that despite the seemingly low risk population (majority white ethnicity, high socioeconomic status, living with partner), a large proportion of new mothers reported symptoms of low mood, anxiety and loneliness during the lockdown. Given the short- and long-term consequences of perinatal mental illness on the physical and psychological well-being of the mother and baby, there is an urgent need for action to support new mothers who have been affected. It is also important to prevent these issues in mothers who are continuing to deliver during different stages of lockdown due to the pandemic.

We found that the perception of how equal the division of chores among household members has been since lockdown began was associated with mental health and coping with the lockdown. Women who felt to a high extent that ‘chores have become more equally divided’ coped better with the situation than those who agreed less strongly with the statement. In our study, 63% of women felt that chores had become more equally divided since lockdown began. This is similar to the results of The Institute for Fiscal Studies [6] which showed that during lockdown, in two parent opposite-gender families, fathers have doubled the time they spend on childcare, but mothers still spent more time on childcare and domestic responsibilities. Unexpectedly, we found that participants who agreed ‘a little’ with the statement ‘chores have become more equally divided’ had better mental health than those who agreed ‘to a high extent‘. It is possible that households with the greatest change in the distribution of chores were those most affected by the lockdown, for example, by the partner losing their job. Nevertheless, there was a dose-response association between the mother receiving support and her feeling able to cope, highlighting the overall importance of support.

Fifty-nine percent of new mothers reported getting enough help with their own health, and this was associated with reduced mental health problems and better coping. In the first year postpartum, it is common for women to experience a wide range of health problems such as tiredness, back pain, urinary incontinence, bowel problems and more frequent coughs, colds, and minor illnesses [15-18]. The association between physical health and mental health problems in the first year after childbirth is an under-researched area, however, a few studies have highlighted the link between physical health problems and mental health [19-21]. Our study provides further evidence that physical health should be an important target for interventions aiming to improve maternal mental health. Additionally, 69% of mothers in our study reported having no or very little time during lockdown to take care of their own health. This emphasises the role of partners, family and social networks in helping mothers postpartum.

We also found that support for mothers in the form of mother-baby or breastfeeding support groups was associated with better mental health (95% CI -.003, .252), while contact with health care professionals was not. This could be due to the type of support received (peer, professional) and the method by which it was given (remote, in-person). Mother and baby support groups may be particularly important for peer support, where other mothers in these groups can relate to the experience of motherhood during lockdown and provide empathetic support and validation [22]; for example, with feelings of being robbed of their experience of motherhood [11, 12]. Health professionals, on the other hand, are likely to provide practical support, and due to COVID-19 restrictions, the majority of the contact with the mothers would have been by phone or online, which makes practical help more difficult. We have previously reported that participants in this survey described their inability to access face-to-face practical support as a reason for changing their infant feeding method [11]. Health visitors have also voiced concerns over the reduced face-to-face home visits during lockdown, with 92% reporting parental mental health as a reason for the concern [Conti & Dow]. Therefore, face-to-face contact may be necessary to provide practical support for the mothers, especially for first-time mothers, whilst online or phone help may be beneficial for emotional and social support.

Additionally, despite the majority of the participants reporting feeling worried (71%), lonely (60%) or down (56%), only 7% had any contact with a mental health professional in the previous week. The lack of care-seeking behaviour among postpartum women was already evident before the lockdown, which has been attributed to stigma associated with mental illness (especially with postpartum depression), normalising symptoms, logistical barriers, and lack of knowledge of available health services [23-25]. Moreover, prior to the pandemic, services in the UK were struggling to meet the demand for mental health services and this is likely to have become more severe with the worsening of the mental health crisis during lockdown [4]. However, it could also be that the participants did not consider their symptoms to be serious enough to seek professional help.

Our study highlighted that travelling to work at least once in the previous week predicted worse mental health outcomes. Among those who travelled for work, the most commonly reported jobs were related to the healthcare sector. Data from previous pandemics and from this pandemic in China and Italy is consistent with this, where 50.3% and 44.6% of healthcare workers reported experiencing depression and anxiety, respectively [26, 27]. The higher risk of mental health problems for frontline workers during the pandemic and for women in the postpartum period in general suggests that these women are at particular risk.

Previous studies have shown that BAME groups, especially Black women, and disadvantaged groups are disproportionately affected by COVID-19 [13, 14]. Our results did not show that ethnicity was associated with mental health or coping, which might be due to the small sample size for the non-white groups, but they indicated that participants whose ability to afford food has been majorly affected, and those earning less than 20-30k a year had significantly worse mental health outcomes. In our study, around a third of participants expressed a minor to major impact of lockdown on their ability to pay rent or mortgage payments, food, and other essentials such as utilities and medication. The quality of the housing, particularly its ability to provide adequate personal and outdoor space for household members, are also important predictors of mental health and vary with socioeconomic status. For example, 12% of households in Great Britain do not have access to a private or shared garden, and Black people in England are four times less likely than white people to have access to outdoor space [28]. These inequalities partially explain the variation in mental health outcomes among different groups. The majority of participants in this study self-identified as white, were married or living with a partner, lived in a house and had access to green space within walking distance. This highlights the need to capture data from a more ethnically and socioeconomically diverse group of women. We were unable to assess the differences in maternal mental health among different ethnic groups due to the underrepresentation of mothers of BAME backgrounds in this survey. Therefore, more effort is being made to include the voices of these mothers.

Data from before the pandemic underlined that mothers of preterm infants experience higher stress, anxiety and depression [29, 30]. The lockdown measures, resulting in a decrease in support networks, changes in hospital policies, and concern about COVID-19 infection, are likely to have exacerbated the mental health problems experienced by those mothers. Our results suggest higher gestational age was associated with better maternal mental health.

Mothers whose babies were born premature, and possibly admitted to the neonatal unit, are at particular risk. Current UK guidance about COVID-19 for parents with babies on the neonatal unit states that parents should have access to their baby and be involved in their care on the neonatal unit [31]. However, access to the neonatal unit and baby is restricted if the parent is showing COVID-19 symptoms. Additionally, after discharge, mothers of premature infants might not be receiving adequate face-to-face practical and emotional support due to COVID-19 restrictions.

Despite the high proportion of new mothers reporting symptoms of low mood, anxiety and loneliness, a high proportion also indicated being able to cope with situation, enjoying the weather, and having the opportunity to chat with friends and family. A review of coping responses towards infectious disease outbreaks showed that coping strategies could take different forms such as problem-solving (seeking alternatives, self- and other-preservation), seeking social support, avoidance, and positive appraisal of the situation [32]. In this study, we investigated going outside for physical activity and relaxation techniques as potential methods used by the participants to cope with the lockdown. A previous study has shown that pregnant women engaging in at least 150 minutes of physical activity during lockdown had lower anxiety and depression scores [10]. We did not find an association between going out for physical activity and maternal mental health; however, direct comparison with the previous study is difficult. The previous study assessed depression and anxiety on formal scales, whereas we examined different symptoms of low mood, anxiety and loneliness.Additionally, 95% of the participants in the present study went outside for a walk or exercise at least once a week, with 62% going at least 4 times per week. These relatively uniform high activity levels reduce our ability to detect an association between variability in activity level and mental health or coping.

In regards to relaxation techniques, 23% of new mothers practiced a form of relaxation at least once a week; more than a third of whom started using these methods during lockdown. The most commonly cited relaxation techniques were yoga, meditation, and breathing exercises. A survey of 5545 Spanish adults during the lockdown found that relaxing activities (such as yoga, gardening and listening to music) were associated with lower anxiety and depression symptoms but only before correction for multiple comparisons [33]. Similarly, in this study, practicing relaxation techniques did not predict coping or mental health, possibly due to the variation in frequency, duration, and type of activities. Given the potential benefit of relaxation techniques and physical activity on anxiety and depression [34-38], further research is warranted.

The main limitations of this study are the cross-sectional design and that the current population is not representative of all new mothers in the UK. Compared to the national data, our participants have higher educational attainment, higher income, are more likely to be married or cohabitating, and are more likely to be of white ethnic background. Further effort is being made to include the experiences of women of BAME backgrounds and from disadvantaged groups to increase the representativeness of our study sample. Another limitation is that we did not formally assess or diagnose depression or anxiety, due to the anonymous nature of the survey which made it not possible to identify participants for follow up. Consequently, we were not able to compare the rates of depression or anxiety in new mothers during lockdown with the rates before the pandemic. However, a larger proportion of the general population do not fit the diagnoses of depression or anxiety, but are rather at risk, which makes our results more generalizable. We will also monitor changes in mental health ‘symptoms’ and coping over the different phases of lockdown.

## Conclusion

The results of this survey indicate that a large proportion of new mothers in the U.K. are experiencing symptoms of low mood, anxiety and loneliness; mothers of preterm infants, with low income, and who are travelling to work are particularly at risk. However, the data also suggests that providing support for mother‘s own health and with household chores are beneficial for maternal mental health and coping. Overall, the findings highlight the urgent need to assess maternal mental health, and to create prevention strategies for mothers who are giving birth during the different stages of lockdown in this COVID-19 pandemic.

## Data Availability

Data will not be publicly available due to ethical approval restrictions.

## Notes

### Competing Interest Statement

The authors declare no conflict of interest with respect to the COVID-19 New Mum Study or this manuscript. MF receives an unrestricted donation for research on infant nutrition from Philips. The remaining authors declare no other conflicts.

### Funding Statement

No funding was received for the survey. All research at Great Ormond Street Hospital NHS Foundation Trust and UCL Great Ormond Street Institute of Child Health is made possible by the NIHR Great Ormond Street Hospital Biomedical Research Centre. The views expressed are those of the author(s) and not necessarily those of the NHS, the NIHR or the Department of Health.

### Author Declarations

Ethical approval was obtained from the UCL Research Ethics committee (0326/017).

